# Temporal Trends in Vaping Use: Demographic Diversity and Healthcare Utilization Outcomes

**DOI:** 10.1101/2020.01.20.20018275

**Authors:** Trevor Nguyen, Teodora Nikova, Eric Luong, Patrick Bottling, Mir Henglin, Elizabeth H. Kim, Florian Rader, Itai Danovitch, Aaron Weinberg, Susan Cheng, Joseph Ebinger

## Abstract

**BACKGROUND:** Use of electronic cigarettes (e-cigarettes) has increased substantially in recent years. Marketed as a safer alternative to conventional tobacco cigarettes, the safety profile of e-cigarettes is now being questioned. Importantly, the clinical consequences of e-cigarette use remain poorly characterized.

**METHODS:** Using data from the electronic health record (EHR), we identified a cohort of e-cigarette users and never-smokers in the region of central Los Angeles. Using information extracted from the EHR, we assessed temporal trends in the demographic characteristics of e-cigarette users, including race/ethnicity and location of residence. We also used data on urgent medical visits (i.e. emergency department encounters and hospital admissions) occurring during the study period (January 2014 to January 2020) and employed a negative binomial regression model to compare healthcare utilization rates between e-cigarette users and never-smokers.

**RESULTS:** We identified a total of 698 e-cigarette users, in addition to 999 never-smokers, with demographic, clinical, and outcomes data available over the duration of the study period. From the beginning to the end of the study period, e-cigarette use increased across all race/ethnicity groups and across all geographic regions of residence. Notably, e-cigarette users experienced a 4.96 higher rate of urgent medical visits when compared to never-smokers evaluated over the same study period.

**CONCLUSION:** E-cigarettes use continues to expand, with evidence of racial/ethnic as well as geographic variation in uptake. Use of e-cigarette devices is associated with substantially higher rates of urgent healthcare utilization. These findings highlight the need for a more granular understanding of e-cigarette use and enhanced attention to the diversity as well as breadth of the population affected by this burgeoning health crisis.

## BACKGROUD

Electronic cigarettes (e-cigarettes) are devices that aerosolize (i.e. vaporize) liquid containing nicotine or other chemicals for inhalation.^1^ Use of these products has rapidly expanded, growing over 46% from years 2014 to 2017, particularly among youth and vulnerable populations.^2-4^ Recent outbreaks of e-cigarette related lung injury have raised concerns about the safety of these products.^5-11^ To examine the diversity of persons at risk and assess the extent to which all users may be prone to developing adverse health outcomes, we assessed temporal trends in e-cigarette use across a large urban medical center based population and examined associated rates of healthcare utilization.

## METHODS

We identified a cohort of e-cigarette users, leveraging electronic health record (EHR) data collected over a period of 6 years (1/1/2014 to 1/15/2020) from the Cedars-Sinai Delivery Network, a high-volume multi-site practice that serves a diverse population of over >300K unique outpatients residing in and around the greater Los Angeles area. Our EHR analytics platform was used to identify presence of e-cigarette use based on both structured and unstructured data. The analysis of unstructured data applies natural language processing along with a collection of concept matching heuristics to identify qualifying records. The applied heuristics are derived from medical ontologies using machine learning techniques, enabling longitudinal assessments to capture duration and frequency of e-cigarette use. E-cigarette use was confirmed via manual chart review performed by trained research staff. To provide comparator data, we also identified a referent group of never-smokers based on discrete data also documented in the EHR. Clinical and demographic data, including residential zip code, for all patients studied were extracted from the EHR, including the number of urgent medical visits occurring during the study period, defined as either emergency department (ED) visits or hospital admissions.

We considered baseline age for each patient as age the start of the study period (1/1/2014). We estimated median household income for patients by linking residential zip code to the 2019 American Community Survey.^12^ We also categorized patients’ residential location using zip code matched to Service Planning Areas (SPAs), which are pre-defined geographic regions that are considered demographically meaningful by the County of Los Angeles.^13^ We collected and analyzed self-reported racial/ethnic data, which was considered missing if the patient refused to answer or the information was not otherwise documented in the EHR.

To compare healthcare utilization rates between e-cigarette users and never-smokers, we constructed a negative binomial regression model adjusting for age, sex, race/ethnicity, and median household income. Follow-up time for e-cigarette users was based the number of days from the first date of recorded substance use to the end of the study (1/15/2020). Follow-up time for non-smokers was calculated as the number of days from the start of the study (1/1/2014) to the end of the study (1/15/2020). All study protocols were approved by the Cedars-Sinai Medical Center institutional review board, with a waiver provided for informed consent.

## RESULTS

We identified N=698 patients with documented e-cigarette use, in addition to a cohort of N=999 randomly selected patients with a documented history of never smoking. Overall, e-cigarette users compared to never-smokers were more likely to be younger in age and female (**Table 1**).

**Table 1.** Demographic characteristics of e-cigarette users and never smokers. We analyzed data extracted from the Cedars-Sinai electronic health record between 1/1/2014 and 1/15/2020. Electronic cigarette use information, including start data of use, were confirmed for all patients via manual chart review. Estimated median household income was determined by linking each patient’s residential zip code to data from the 2012 American Census Survey.

| **Characteristics** | **E-Cigarette Users**<br>**(N=698)** | **Never-Smokers**<br>**(N=999)** |
| --- | --- | --- |
| Age in years, mean (SD) | 39.2 (16.4) | 52.1 (18.9) |
| Female, n (%) | 272 (39.0%) | 632 (63.3%) |
| Race, n (%) |  |  |
| White | 511 (73.2%) | 766 (76.7%) |
| African American | 61 (8.7%) | 150 (15.0%) |
| Asian | 48 (6.9%) | 46 (4.6%) |
| Native Hawaiian/Pacific Islander | 4 (0.6%) | 0 |
| American Indian/Alaska Native | 1 (0.1%) | 2 (0.2%) |
| Missing | 38 (5.4%) | 12 (1.2%) |
| Other | 35 (5.0%) | 23 (2.3%) |
| Household Income, est. median (SD) | \$73,806 (\$28,719) | \$85,001 (\$35,581) |

Among all e-cigarette users, racial minorities represented 21% of the population by the end of the study period (**Figure 1**). Asians and Native Hawaiian/Pacific Islanders were more likely to be e-cigarette users than never-smokers in our sample; by contrast, African Americans were more likely to be never-smokers (**Table 1**).

**Figure 1.**
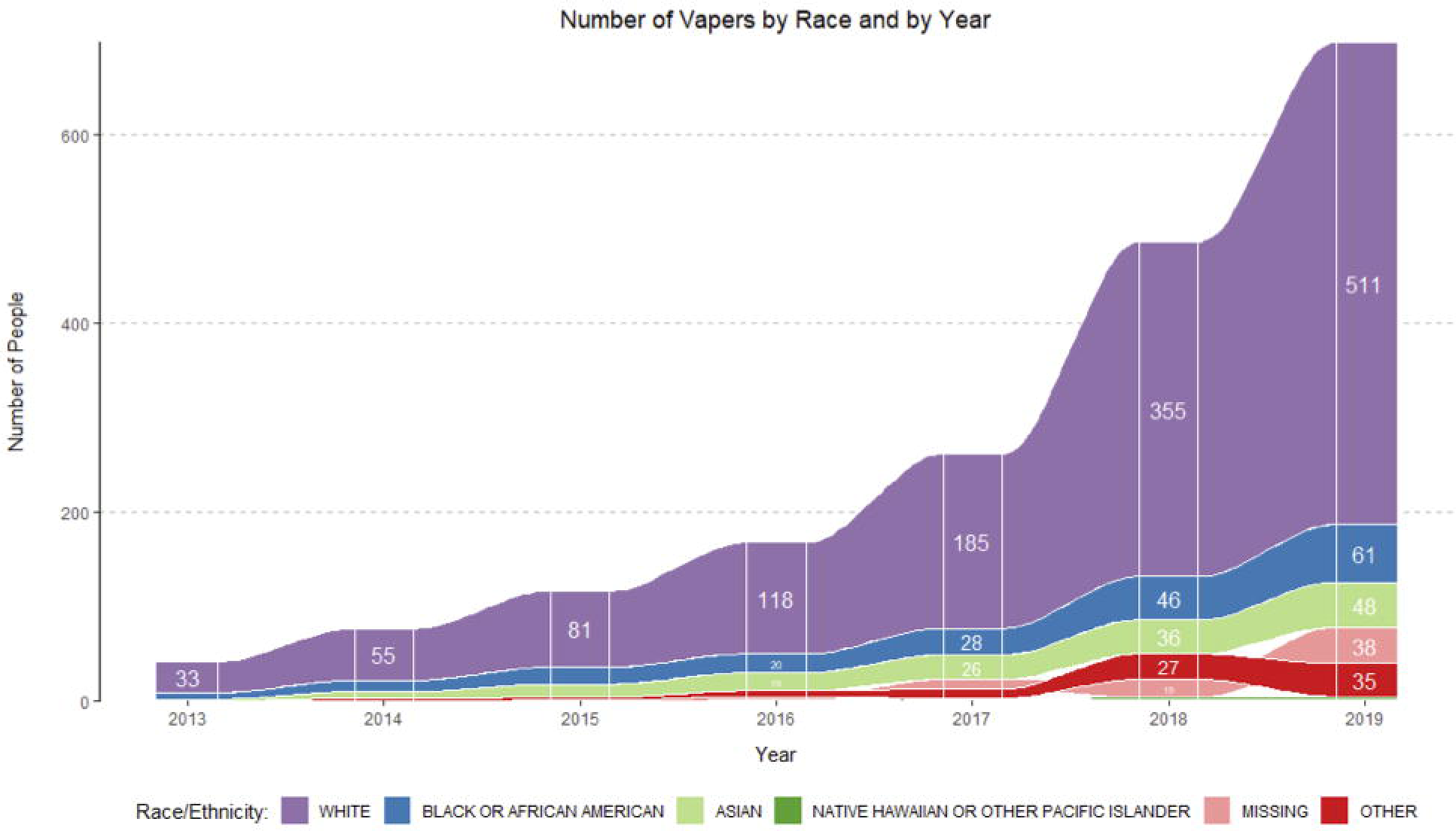
Prevalence of e-cigarette users by racial/ethnic group per year. We analyzed data extracted from the Cedars-Sinai electronic health record between 1/1/2014 and 1/15/2020. Electronic cigarette use information, including start data of use, were confirmed for all patients via manual chart review. All data were categorized by self-reported race/ethnicity group, as documented in the medical record.

With respect to regional diversity, we observed an increase in e-cigarette use across all SPAs during the study period. Starting in 2017, an increased rate of use was seen particularly in SPA 5 (**Figure 2**).

**Figure 2.**
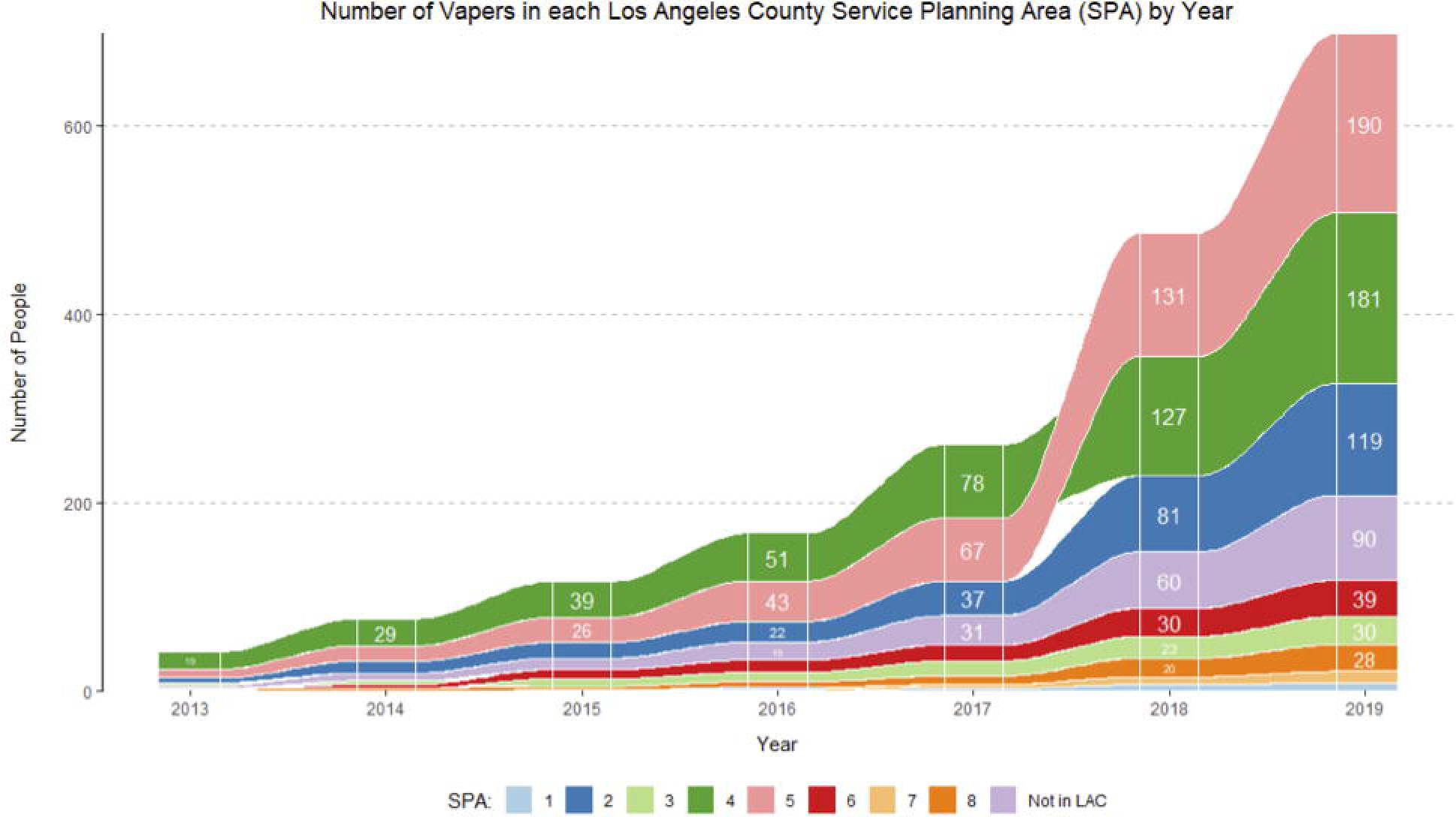
Prevalence of e-cigarette users per year by region. We analyzed data extracted from the Cedars-Sinai electronic health record between 1/1/2014 and 1/15/2020. Electronic cigarette use information, including start data of use, were confirmed for all patients via manual chart review. Patient region of residence, based on service planning area (SPA), was determined by linking residential zip codes to the Los Angeles County SPA database key.

Overall, e-cigarette users experienced a 4.96 higher rate of urgent medical visits (i.e. emergency department visits and hospital admissions) when compared to never-smokers examined during the same study period.

## DISCUSSION

We observed that use of e-cigarettes is rapidly increasing over time, across the greater Los Angeles and surrounding areas, with evidence of racial/ethnic as well as geographic variation in uptake. Importantly, we also found that use of e-cigarette devices is associated with a substantially elevated rate of urgent healthcare utilization. These findings highlight the need for a more granular understanding of e-cigarette use at the community level and enhanced health attention to the diversity as well as breadth of the population affected by this burgeoning health crisis.

Defined geographic areas in Los Angeles, SPAs are utilized by public health officials to plan clinical services based on community needs.^13^ We found an increase in e-cigarette use in all SPAs, with a rapid increase in SPAs 4 and 5 starting in 2017. This likely reflects the importance of socioeconomic and community factors that underly e-cigarette use, including proximity to shops that sell the devices and advertising to specific populations.^14-17^ To date, our limited understanding of e-cigarette use patterns and health risk remain dependent large national surveys which often lack either longitudinal follow-up, temporal relationships between health events and granular location data, *limiting efforts to assess geographic and* 36 *social disparities*.^18-21^ Our findings highlight that e-cigarette use rates are not changing equally in all communities and that efforts to curtail use may require locally tailored solutions.

Of particular concern is the significantly higher rate of urgent healthcare utilization among e-cigarette users compared with never-smokers. While this study was unable to control for pre-existing comorbid conditions, which may predispose to excess healthcare utilization, extensive evidence points to the health risks of e-cigarette use which may drive healthcare utilization. Specifically, numerous studies demonstrate that compounds found in e-cigarette vapor, such as Acrolein, cause adverse cardiovascular effects including reduced myocardial contractility, increased thrombosis risk and induction of vascular injury.^22-26^ Analyses of the National Health Interview Survey and Population Assessment of Tobacco and Health datasets demonstrate a positive association between e-cigarettes and risk of myocardial infarction; Behavioral Risk Factor Surveillance System data now clearly links e-cigarettes with an increased risk of any form of cardiovascular disease, a risk which is multiplied when these devices are used in combination with traditional cigarettes.^27-29^ Our study suggests that these adverse effects are not necessarily rare events, but rather are driving patients to seek urgent medical attention at markedly elevated rates.

## Data Availability

Deidentified data will be made available upon request, in accordance with current standards.

